# Building Back Better after COVID-19: a systematic scoping review of wicked problems affecting developed countries and implications for global governance

**DOI:** 10.1101/2021.04.26.21256126

**Authors:** Robin van Kessel, Brian Li Han Wong

## Abstract

The COVID-19 pandemic is a textbook example of a ‘wicked problem’, one which is complex, open-ended, unpredictable, or intractable and seems resistant to any solution. This presents a window of opportunity to explore other wicked problems and their implications after the pandemic. A systematic scoping review was conducted to investigate the COVID-19 aftermath and identify public health topics which may be of great significance in the years to come. Through the adoption of three megadrivers as fundamental drivers of change (globalisation, demographic change, and digitalisation), it narratively explored how different wicked problems – and the driving mechanisms which sustain them – persist. It further explored the implications of these public health topics on global (health) governance. While the wicked problems mapped in this article show a large variance in where their apparent roots lie, they share one factor in common: health. These wicked problems must be first and foremost addressed if we as a globalised world are to successfully and sustainably build back better from COVID-19.

**Summary Box:** *What is already known?:*

- COVID-19 is a textbook example of a wicked problem; a problem that is complex, open-ended, unpredictable, or intractable and seems resistant to any solution;
- Digitalisation, globalisation, and demographic change are seen as the three megadrivers of change and are theorised to create and sustain modern wicked problems;
- The megadrivers respectively have complex relationships with health; this includes both positive and negative associations.

*What are the new findings?:*

- A series of wicked problems exist in multiple domains of society that continue to obstruct the progress of the 2030 Agenda for Sustainable Development;
- These wicked problems each have roots in the three megadrivers: digitalisation, globalisation, and demographic change;
- There exists a complex interrelation between the megadrivers, wicked problems, and health.

*What do the new findings imply?:*

- It currently remains unclear to what extent wicked problems and megadrivers respectively affect health outcomes; therefore, further research is indicated;
- A ‘once in a generation’ opportunity has presented itself to build back better from the COVID-19 pandemic by addressing the existing wicked problems; therefore, governance mechanisms should follow and adapt accordingly;
- Wicked problems have bidirectional implications for modern policy; this has created an environment for wicked problems to manifest and sustain themselves, which in turn produce further policies that sustain wicked problems.

## Introduction

“Health is a political choice and politics is a continuous struggle for power among competing interests” [1]. In this article, Kickbusch highlights how health is inherently political and part of a complex, interrelated, and interdependent system [1,2]; meaning it requires extensive knowledge of the other factors, as well as key stakeholders and their (political) priorities to be able to instigate and influence change. More specifically, health is shaped on a political level by factors like the distribution of power and resources, at local, national, and global levels respectively and these factors can only be influenced in domains other than health [3]. In terms of resources, this does not only refer to the distribution of money. It also includes human resources and the ill-preparedness of the public health workforce to tackle problems like COVID-19 due to the erosion of public health systems [4]. On top of that, COVID-19 has permeated into other dimensions of society, for instance: technology, politics, domestic and international governance, and globalisation [5,6].

The COVID-19 pandemic is therefore a text-book example of a ‘wicked problem’: a problem that is complex, open-ended, unpredictable, or intractable and seems resistant to any solution [7]. While COVID-19 is a crucial problem to solve, it is important to be aware that many wicked problems exist concurrently alongside COVID-19 (e.g. climate change, humanitarian crises and conflicts, digital divide). Some wicked problems are connected to the pandemic as well, such as vaccine hesitancy and vaccine diplomacy, that is the branch of global health diplomacy that concerns the use or delivery of vaccines [8], such as coercively using vaccines to gain influence on the global scale--a recurring phenomenon during large outbreaks of disease, previously seen during the smallpox outbreaks [9].

With the conditional market authorisation of various vaccines for COVID-19 in the European Union—a model for the developed world—a window of opportunity has arisen to explore other wicked problems and their implications after the pandemic. The United Nations and OECD point out that this is a ‘once in a generation’ opportunity to build back better [10,11]; to use the shock that COVID-19 caused to society in order to address complex/wicked problems that threaten the 2030 Agenda for Sustainable Development. By investigating current wicked problems in the domains of sociology, political science, education, economy, and ecology, this scoping review aims to contribute to investigating the COVID-19 aftermath and to identify public health topics which may be of great significance in the years to come. Subsequently, we explore what the implications of these public health topics are on global (health) governance.

## Methods

A scoping review is particularly useful when a body of literature has a complex and heterogeneous nature and is previously under-researched [12,13]. Research on what the ‘new normal’ will be and how society will function/’build back better’ post-pandemic is still in its infancy, making this methodology a fitting option to further understand this topic. This article adhered to the systematic nature of the PRISMA 2020 framework by establishing clear eligibility criteria, defining information sources, reporting the complete search query, and outlining the data collection process and the data items that were looked for [14]. The assessment of bias was not translated into this methodology, as it is not within the scope of a systematic scoping review [13]. Findings were reported based on the PRISMA 2020 framework by stating the exact number of identified articles before and after screening, providing an overview of the contents of the individual articles, and a synthesis of the included articles [14].

### Search Strategy and Selection Criteria

A search strategy was devised to search the following databases: SocINDEX (sociology), Web of Science (political science), Educational Resource Information Center (education), GreenFILE (ecology), and EconLit and Business Source Complete (economics). Databases were accessed through EBSCO Host. The exact search terms for each academic database are added in eTable 1 in the Supplementary Materials.

**Table 1.** Key characteristics of the included studies.

| Publication | Type of Article | Field | Wicked Problem |
| --- | --- | --- | --- |
| Scherer & Voegtlin, 2020[18] | Symposium | Governance | Participative/reflexive governance has to find a balance between (1) participation and expertise; (2) diversity and consensus; and (3) polycentricity and centralisation. |
| Thomson & Ip, 2020[19] | Editorial | Political Science | Political systems worldwide are increasingly resorting to excessive and disproportionate emergency measures that spell grave dangers for civil rights and liberties. It risks the creation of a new pandemic after COVID-19: one of authoritarian rule. |
| Brammer, Branickim, & Liennenluecke, 2020[20] | Article | Sociology | Societalisation: a macro-cultural concept concerned with the process through which a society shifts in its collective perspective on specific problems by experiencing a revelatory crisis or upheaval. |
| El-Hani & Machado, 2020[21] | Viewpoint | Sociology | Building an integrated view, both trans- and interdisciplinary, suffers from epistemological challenges due to differences in stakeholder perspectives, values, interests, and knowledge. |
| Samuel et al., 2020[22] | Article | Public Health, Economy | As a result of the pandemic, collective feelings as fear, despair, anguish, disconnection, and isolation have perpetuated society, which indicate complex mental healthcare challenges down the road.<br>The pandemic also caused substantial global disruptions, resulting in trillions of euros in estimated losses and many (smaller) businesses and even entire industries being forced to close down. Unemployment can be directly and indirectly affected by this development: directly as businesses that shut down for good let go of their workforce; indirectly as businesses that try to survive look at cost-containment. |
| Drake & Reid, 2020[23] | Article | Education | Current education systems that teach traditional problem-solving skills prevent wicked problems from being addressed by future generations. Traditional problem-solving does not or in a very limited way allow for multilayered complexities and big-picture perspectives, inherently limiting the solutions that can be imagined. |
| Gardiner, 2020[24] | Article | Education | Education is currently fundamentally limited by their approach to knowledge. Currently, education is still focused on acquiring knowledge, rather than understanding the limits, benefits, and nuances of the knowledge possessed. As wicked problems become more prevalent, simply “knowing” things will not help to address them. |
| King, 2021[25] | Article | Political Science | Brexit is interpreted as a wicked problem that stems from other wicked problems, such as the 2008 banking crisis and decades of inner commotion in the Conservative Party over the issue of Europe linked with a sentiment of a ‘British identity’ that led to a particularly devastating (social) medial campaign. The article is retrospective in nature, but highlights problems that other countries either face right now or run the risk of facing in the near future. |
| Pietrocola et al., 2020[26] | Article | Education | COVID-19 is framed as a manufactured problem of great complexity that people that have been through the current education system are unable to solve due to the fact they were never familiarised with problems of this complexity or magnitude. This then serves as a contributing factor as to why problems like climate change, forest fires, global warming, environmental issues, and pandemics are very difficult to conceptualise, let alone tackle. |
| Fuentes et al., 2020[27] | Article | Ecology | Climate Change and global warming are caused by rising levels of greenhouse gasses, irrespective of the geographical location of their emission. As pollution comes from a wide array of places, it is particularly difficult to address. Additionally, the information asymmetry surrounding climate change further inhibits the ability to address the problem. |
| Gras et al., 2020[28] | Article | Problem-solving, Business | Use of reductive tendency in problem solving: complex problems are often simplified into cognitively manageable parts. While the process has its benefits, the main downside is that it ignores the intricacies of a complex problem, making it even more difficult to work towards a real solution. The solutions that are crafted are based on the notion a problem is less complex than it actually is and that that problem can be solved, even though the actual requirements for solving the problem are absent. |
| World Health Organization, 2020[29] | Report | Ecology, Health Protection, Sustainability, Urban Planning | (1) Human pressures on the natural environment, ranging from deforestation, intensive pollution of agricultural practices, and extensive consumption of wildlife;<br>(2) Antimicrobial-resistant pathogens becoming more prevalent; management of sources where these pathogens reside is vital;<br>(3) More than 90% of people worldwide breathe polluted air that exceeds WHO air quality guidelines, which is largely caused by the burning of fossil<br>fuels that also drive climate change;<br>(4) Disease caused by lack of access to food or excessive intake of unhealthy food forms the largest cause of global ill-health and there is an established need for a quick transition to sustainable, healthy, and nutritious diets;<br>(5) Currently, cities worldwide are responsible for 60% of greenhouse gas emissions, which can be addressed by improvements in urban planning and infrastructure;<br>(6) Fossil fuels continue to be subsidised by governmental programs. |
| Smit et al., 2020[30] | Report | Workforce, Economy | The report outlines three elements that compile the wicked problem of “labour market mismatch”<br>(1) Europe is an amalgamation of highly varied local labor markets that have become more geographically concentrated over time;<br>(2) A potential shortage of skilled workers may be experienced once the economy recovers due to a combination of ageing and automation;<br>(3) Skill requirements of Europe’s workforce are likely to change due to automation. |
| Ansell, Sørensen, & Torfing, 2020[31] | Article | Public Administration, Leadership | A zero-error culture is still common across public administrations, indicating that mistakes are not allowed to be made. Errors are still portrayed as “the end of the world and evidence of incompetence or lack of motivation,” rather than a first step towards a new solution.<br>Public administrations still rely heavily on control, rather than trust, which inhibits flexibility, creativity, and adaptation.<br>Leadership needs to be remodeled, moving from the traditional “principal” role towards a role of “steward” in which followers are encouraged, enriched, and involved in working collaboratively towards a solution. Leaders also have to learn to operate in highly unpredictable circumstances and try to solve issues without knowing all the details. |

Table 1. Key characteristics of the included studies.
| Publication | Type of Article | Field | Wicked Problem |
| --- | --- | --- | --- |
| Scherer & Voegtlin, 2020[19] | Symposium | Governance | Participative/reflexive governance has to find a balance between (1) participation and expertise; (2) diversity and consensus; and (3) polycentricity and centralisation. |
| Thomson & Ip, 2020[20] | Editorial | Political Science | Political systems worldwide are increasingly resorting to excessive and disproportionate emergency measures that spell grave dangers for civil rights and liberties. It risks the creation of a new pandemic after COVID-19: one of authoritarian rule. |
| Brammer, Branickim, & Liennenluecke, 2020[21] | Article | Sociology | Societalisation: a macro-cultural concept concerned with the process through which a society shifts in its collective perspective on specific problems by experiencing a revelatory crisis or upheaval. |
| El-Hani & Machado, 2020[22] | Viewpoint | Sociology | Building an integrated view, both trans- and interdisciplinary, suffers from epistemological challenges due to differences in stakeholder perspectives, values, interests, and knowledge. |
| Samuel et al., 2020[23] | Article | Public Health, Economy | As a result of the pandemic, collective feelings as fear, despair, anguish, disconnection, and isolation have perpetuated society, which indicate complex mental healthcare challenges down the road.<br>The pandemic also caused substantial global disruptions, resulting in trillions of euros in estimated losses and many (smaller) businesses and even entire industries being forced to close down. Unemployment can be directly and indirectly affected by this development: directly as businesses that shut down for good let go of their workforce; indirectly as businesses that try to survive look at cost-containment. |
| Drake & Reid, 2020[24] | Article | Education | Current education systems that teach traditional problem-solving skills prevent wicked problems from being addressed by future generations. Traditional problem-solving does not or in a very limited way allow for multilayered complexities and big-picture perspectives, inherently limiting the solutions that can be imagined. |
| Gardiner, 2020[25] | Article | Education | Education is currently fundamentally limited by their approach to knowledge. Currently, education is still focused on acquiring knowledge, rather than understanding the limits, benefits, and nuances of the knowledge possessed. As wicked problems become more prevalent, simply “knowing” things will not help to address them. |
| King, 2021[26] | Article | Political Science | Brexit is interpreted as a wicked problem that stems from other wicked problems, such as the 2008 banking crisis and decades of inner commotion in the Conservative Party over the issue of Europe linked with a sentiment of a ‘British identity’ that led to a particularly devastating (social) medial campaign. The article is retrospective in nature, but highlights problems that other countries either face right now or run the risk of facing in the near future. |
| Pietrocola et al., 2020[27] | Article | Education | COVID-19 is framed as a manufactured problem of great complexity that people that have been through the current education system are unable to solve due to the fact they were never familiarised with problems of this complexity or magnitude. This then serves as a contributing factor as to why problems like climate change, forest fires, global warming, environmental issues, and pandemics are very difficult to conceptualise, let alone tackle. |
| Fuentes et al., 2020[28] | Article | Ecology | Climate Change and global warming are caused by rising levels of greenhouse gasses, irrespective of the geographical location of their emission. As pollution comes from a wide array of places, it is particularly difficult to address. Additionally, the information asymmetry surrounding climate change further inhibits the ability to address the problem. |
| Gras et al., 2020[29] | Article | Problem-solving, Business | Use of reductive tendency in problem solving: complex problems are often simplified into cognitively manageable parts. While the process has its benefits, the main downside is that it ignores the intricacies of a complex problem, making it even more difficult to work towards a real solution. The solutions that are crafted are based on the notion a problem is less complex than it actually is and that that problem can be solved, even though the actual requirements for solving the problem are absent. |
| World Health Organization, 2020[30] | Report | Ecology, Health Protection, Sustainability, Urban Planning | (1) Human pressures on the natural environment, ranging from deforestation, intensive pollution of agricultural practices, and extensive consumption of wildlife;<br>(2) Antimicrobial-resistant pathogens becoming more prevalent; management of sources where these pathogens reside is vital;<br>(3) More than 90% of people worldwide breathe polluted air that exceeds WHO air quality guidelines, which is largely caused by the burning of fossil fuels that also drive climate change;<br>(4) Disease caused by lack of access to food or excessive intake of unhealthy<br>food forms the largest cause of global ill-health and there is an established need for a quick transition to sustainable, healthy, and nutritious diets;<br>(5) Currently, cities worldwide are responsible for 60% of greenhouse gas emissions, which can be addressed by improvements in urban planning and infrastructure;<br>(6) Fossil fuels continue to be subsidised by governmental programs. |
| Smit et al., 2020[31] | Report | Workforce, Economy | The report outlines three elements that compile the wicked problem of “labour market mismatch”<br>(1) Europe is an amalgamation of highly varied local labor markets that have become more geographically concentrated over time;<br>(2) A potential shortage of skilled workers may be experienced once the economy recovers due to a combination of ageing and automation;<br>(3) Skill requirements of Europe’s workforce are likely to change due to automation. |
| Ansell, Sørensen, & Torfing, 2020[32] | Article | Public Administration, Leadership | A zero-error culture is still common across public administrations, indicating that mistakes are not allowed to be made. Errors are still portrayed as “the end of the world and evidence of incompetence or lack of motivation,” rather than a first step towards a new solution.<br>Public administrations still rely heavily on control, rather than trust, which inhibits flexibility, creativity, and adaptation.<br>Leadership needs to be remodeled, moving from the traditional “principal” role towards a role of “steward” in which followers are encouraged, enriched, and involved in working collaboratively towards a solution. Leaders also have to learn to operate in highly unpredictable circumstances and try to solve issues without knowing all the details. |

Documents had to be published in 2020, discuss wicked problems and their potential implications, be published in English, focus on wicked problems in the developed world, and be academic in nature (meaning entries in scientific journals). Reports from large international organisations like the World Health Organization or McKinsey Global Institute were also eligible. No limitation was put on target population.

### Data Analysis

The conceptual model by Mehrolhasani and colleagues takes a societal focal point, acknowledging that each society has a history and moves towards a possible, probable, preferable, or desirable future [15], while recognising that governance heavily impacts the development of societies and what future is considered desirable. The model is applied retrospectively in this article in order to explore under what circumstances the current environment that hosts the present wicked problems was deemed a preferable or desirable future. For governance, the TAPIC framework is utilised, which divides governance in five domains: transparency, accountability, participation, integrity, and capacity [16].

This article adopts the three megatrends by Petersen and Steiner as fundamental drivers of change [17]: (1) globalisation; (2) demographic change; and (3) digitalisation. Globalisation, in this context, is conceptualised as “the increasing economic, political, social and cultural integration of countries and people” (p.12); demographic change refers to “a change in the size and structure of the population” (p.16); and digitalisation refers to “the global expansion of information and communication technologies, also including networking and acceleration tendencies that cause considerable changes in the political, social, cultural, and economic structures of societies” (p.20). Using these three megatrends, we narratively explore how different wicked problems may persist, which grants more insight in the driving mechanisms that sustain them. Nevertheless, we also have to consider that the megadrivers have their individual impact on health, creating a complex conceptual model (see Figure 1). Further information on the influence of the megadrivers on health is added in the eMethods of the Supplementary Material.

**Figure 1.**
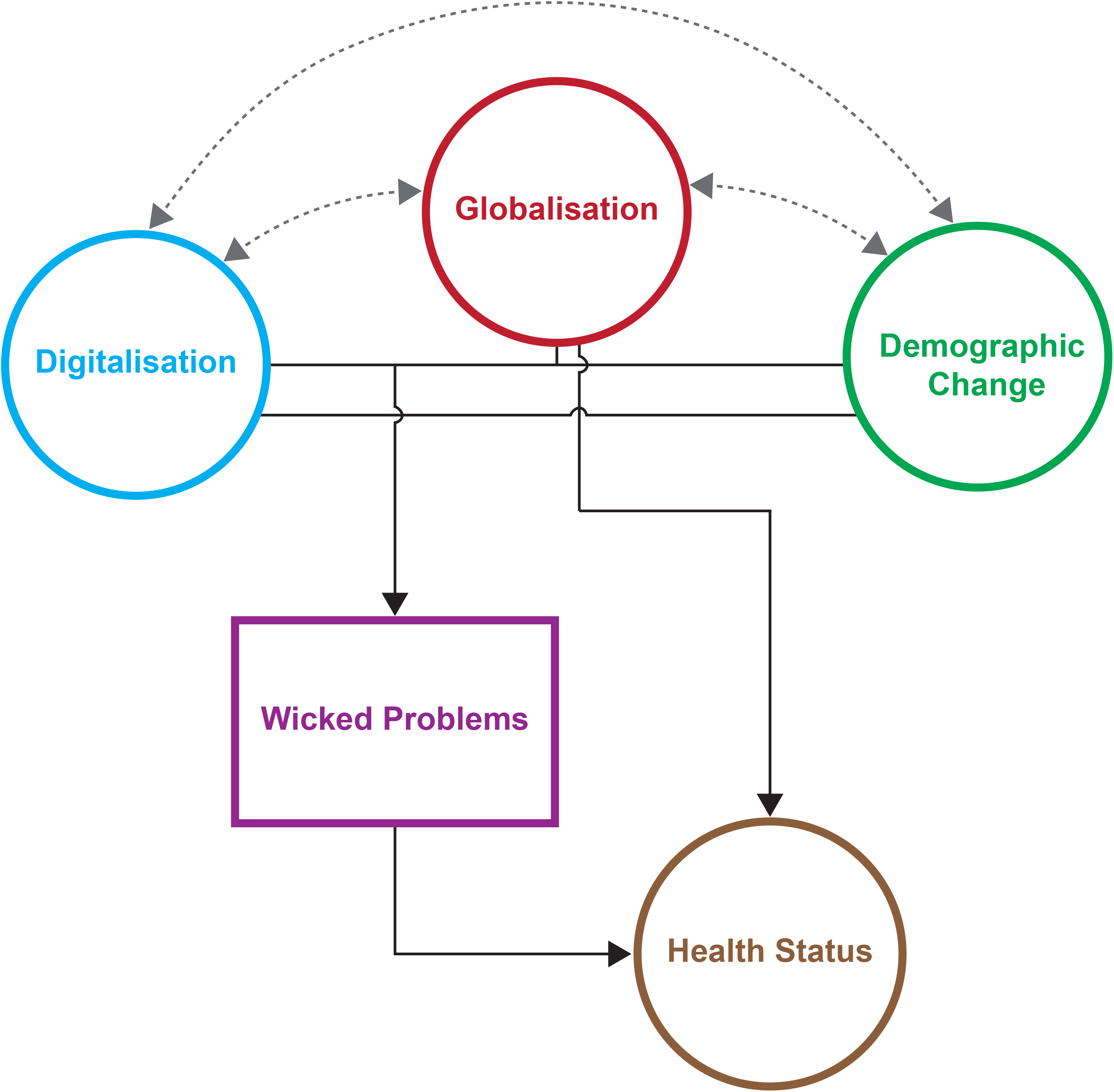
The interrelationships between the megadrivers, wicked problems, and health outcomes.

**Figure 2.**
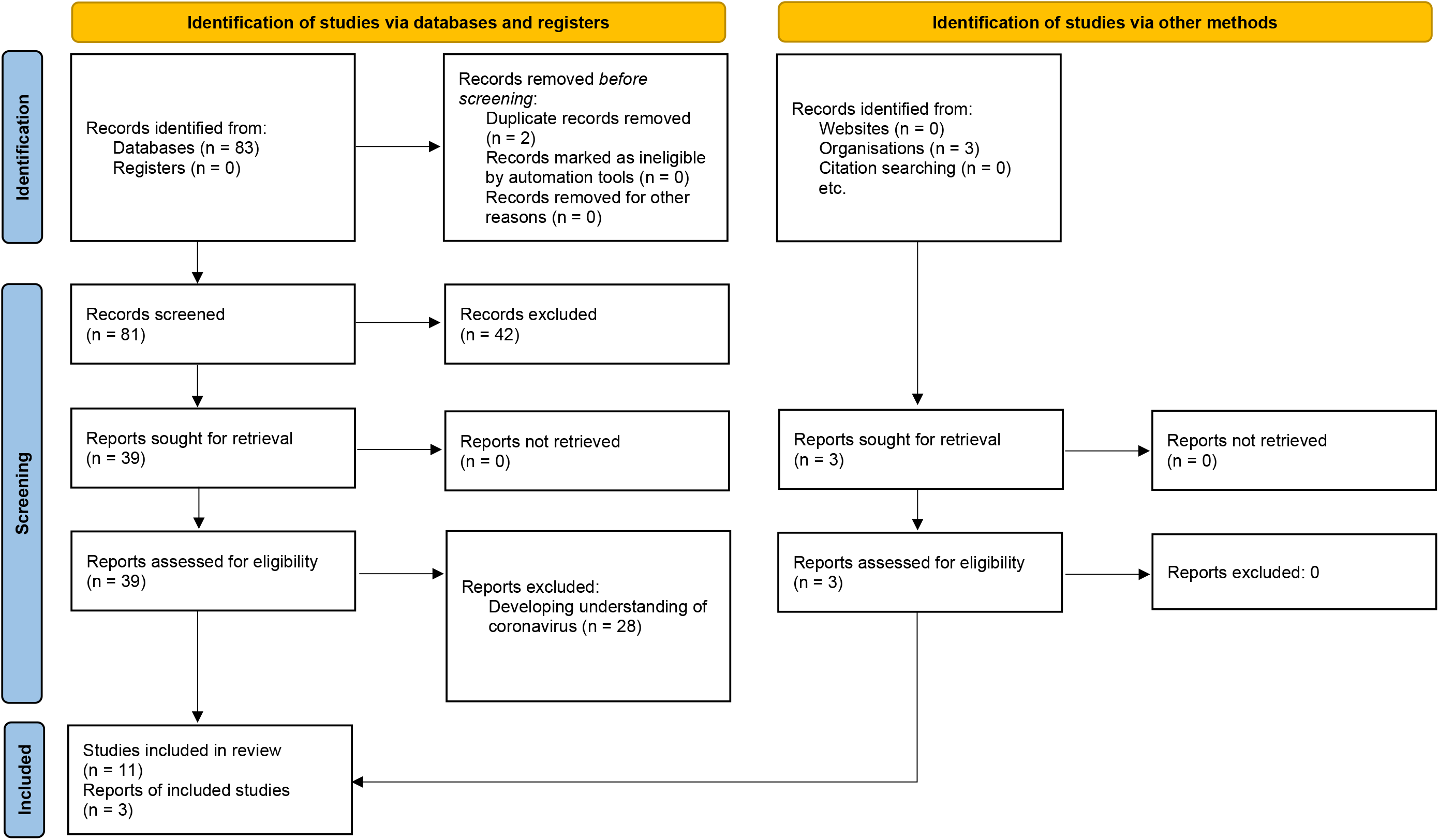
The PRISMA flowchart showcasing the data collection process.

## Results

Academic database searches yielded 83 results and 3 reports from organisations were identified. After removing two duplicates, 84 results were explored based on the eligibility criteria. In the end, 14 documents were included in this study. Figure 1 illustrates the selection process through a PRISMA 2020 flowchart. Key characteristics of the individual articles are included in Table 1, along with a description of the wicked problem described in that article.

When comparing these wicked problems to the megatrends, a reflection of all three can be found in most of the wicked problems. More specifically, the availability of information as a result of digitalisation is a recurring theme among the problems. Information that was previously only limitedly available is now increasingly and more readily available in online formats. This places greater emphasis on the need to gather, analyse, and understand the nuances of information or data. Metadata can be considered an up-and-coming field that shifts the status-quo of how to approach learning and information processing. Failure to understand the nuances, origins, and potential limitations and biases that information and its sources have, can contribute to the devastating effects of dis- and misinformation that we are currently experiencing in the COVID-19 pandemic [32].

Globalisation also plays an increasing role as populations, economies, cultures, and political systems of different countries keep interacting with each other. It inherently breaks down the concepts of a “national identity” as people and cultures continue to mix, which is prone to backlash—as seen in the situation of Brexit [25]. In times of crisis, globalisation is also able to incentivise authoritarian measures as crisis responses, especially if they appear to work [19]. A major problem here is that translation of policies from one setting to another is a complex process in its own right [33]. Failure to account for the complexity of translating policies could result in diminished effectiveness and lower adherence to policy. Especially in times of crises and when tackling wicked problems, it is paramount that potential policies/solutions resonate with implementers and those for whom the policies/solutions are designed.

Wicked problems can affect different population groups in different ways. COVID-19 has been shown to disproportionately negatively affect people over the age of 60 compared to people under 60 [34]. People over 50 were found to be more susceptible to believe misinformation campaigns [35]. An explanation for this susceptibility is that older adults tend to be newcomers in digital environments and therefore may not have the same levels of digital literacy compared to younger population groups. Also, certain social changes play a role in the increased susceptibility, namely an increase in interpersonal trust and more difficulty with detecting lies [36–38].

## Discussion

The aim of this study was to explore current wicked problems through the lenses of three megadrivers of change, identify the effect they may have on public health, and investigate the implications these public health topics can have on global (health) governance.

The wicked problems mapped in this article show a large variance in where their apparent roots lie. Nevertheless, they share one common factor among all of them, namely their relation to health. Using an updated version of the 1991 Dahlgren and Whitehead model of social determinants of health that has integrated the role of information technology [39], the wicked problems can individually be connected to potential health effects. The problems seem to have manifested predominantly in the layers beyond individual health (e.g. social circles and communities; education and employment; general socio-economic, cultural, and environmental conditions; and communication technologies). That being said, the wicked problems are not only individually connected to certain effects on health, but can also affect each other. A particularly clear example of this is the problem of education teaching problem-solving skills that are not able to address modern-day problems, which then perpetuates an inability of society to address wicked problems or the pandemic-caused lockdown creating the conditions for domestic violence to rise in prevalence [40].

The model by Mehrolhasani can provide an explanation for the presence of wicked problems on these societal levels and their absence on the individual level [15]. Retrospectively, a society was likely desired in which individuals were raised and trained to take care of their own problems directly. However, larger-scaled problems, such as the mapped wicked problems, were poorly addressed or even conceptualised. This is reflected in the presence of wicked problems that pertain to education, which explain that problem-solving skills are taught in a traditional or even archaic way that does not correspond anymore with the modern problems at hand [24,26]. When subsequently looking through a governance lens, the TAPIC framework can explain why these problems manifested in these respective layers of society and why wicked problems are barely present on the individual level. On an individual level, these domains can be influenced by a single person or a small, coordinated group. However, as problems start to become larger, harder to conceptualise, and require more people, coordination, and collaboration, addressing them becomes increasingly more difficult as one of the five TAPIC dimensions is likely to fall short.

The strong focus on building back better requires a proper understanding of the problems that were ingrained in the former status quo. This article furthers this understanding by mapping wicked problems through systematic searches and data collection. Wicked problems were connected to large societal processes or megadrivers that contribute to their sustenance. Afterwards, the implications for governance were discussed as a whole, as well as per megadriver specifically, leading to an explanation as to why the wicked problems manifested in these particular societal layers. This discussion also gives rise to potential avenues that can be explored to improve or explore new forms of governance—especially using the rich influence of the megadrivers to propel this change forward.

There are some limitations that have to be considered. Firstly, the data provided in this article is meant to incentivise further and deeper analysis and the results should be interpreted as such. Even though we used a clear and transparent methodology to produce the findings, the lack of quality assessment and systematic synthesis limit the use of this article beyond sensitising, scoping, informing, and incentivising. Secondly, some wicked problems may not yet be classified as such in modern literature. As such, these problems are not covered by this article.

In order for the opportunities presented by digitalisation and digital transformations to be maximized (see also Holly, Wong, et al., 2021; forthcoming), policymakers and other stakeholders must invest in: improving access to digital platforms, affordable broadband, and appropriate devices; the inclusion of vulnerable populations (in particular youth [aged 15-24; {40}], who are often the driving force behind digital uptake and advancement); building and improving digital capacity (e.g. skills and literacy), not only among professionals and stakeholders, but across the population [42]; and (re)building public trust in the digital world and information systems after the devastating utilisation during the COVID-19 pandemic (also with regards to the future developments of AI and the prominent role it is likely going to play in society). With regards to globalisation, stakeholders and change agents should consider a new social contract—one that is more geared towards community-based approaches and pooling risks and resources [43]. The change in demographic can further push a new social contract, as older people can contribute their experience on where the old social contract fell short, while the innovativeness of young people can be utilised to form the beginnings of a new social contract. This is also pushed by the WHO-UNICEF-Lancet Commission, who call for children and youth to be put at the heart of the concepts of sustainability [44], recognising that these children will be the change agents that have to address global and wicked problems in the future.

Ultimately, these suggestions are only the starting point in the road to address the wicked problems mapped in this article. The recent adoption of General Comment 25 of the UN Convention of the Rights of the Child is a milestone accomplishment, one of many to come on the road to building back better from COVID-19. As a globalised society, this once-in-a-lifetime opportunity to address the structural issues which have been present for decades is one that we can ill afford to miss.

## Data Availability

Since it is a review article, all data is available on scientific databases

## Acknowledgement

We would like to thank Prof Helmut Brand and Dr Timo Clemens for their thought-provoking discussions during the writing process.

## Notes

### Competing Interest Statement

The authors have declared no competing interest.

### Funding Statement

No funding was acquired for this article.

### Author Declarations

All ethical guidelines have been followed.

