## Supplementary Materials for "Building Back Better after COVID-19: a systematic scoping review of wicked problems affecting developed countries and implications for global governance"

**eTable 1.** An overview of the search queries per academic database.

**eMethods.** The effects of megadrivers on health.

**eReferences**

This supplementary material has been provided by the authors to give readers additional information about their work.

eTable 1. An overview of the search queries per academic database.

| **Database** | **Search Query** | **Hits** |
| --- | --- | --- |
| Web of Science | TOPIC: ("wicked problem*" OR "complex problem*" OR "turbulent problem*" OR "unsolvable problem*" OR "wicked challenge*" OR "complex challenge*" OR "divergent problem*") AND TOPIC: ("coronavirus*" OR "covid*" OR "2019-ncov" OR "2019ncov" OR "severe acute respiratory syndrome coronavirus 2" OR "post-acute COVID-19 syndrome" OR "COVID-19 vaccin*" OR "COVID19 vaccin*")  Timespan: 2020-2021. Indexes: SCI-EXPANDED, SSCI, A&HCI, CPCI-S, CPCI-SSH, ESCI. | 55 |
| ERIC, EconLit, Business Source Complete, and SocIndex^1^ | (“wicked problem*” OR “complex problem*” OR “turbulent problem*” OR “unsolvable problem*” OR “wicked challenge*” OR “complex challenge*” OR “divergent problem*”) AND (“coronavirus*” OR “covid*” OR “2019-ncov” OR “2019ncov” OR “severe acute respiratory syndrome coronavirus 2” OR “post-acute COVID-19 syndrome” OR “COVID-19 vaccin*” OR “COVID19 vaccin*”)  Limiters - Scholarly (Peer Reviewed) Journals; Publication Type: Academic Journal, Dissertation; Document Type: Article, Editorial, Report; Language: English; Publication Type: Dissertation, Journal Article; Publication Type: Dissertations/Theses (All), ERIC Publications, Journal Articles; Language: English; Publication Type: Academic Journal; Document Type: Article, Editorial, Report; Document Type: Article, Dissertation, Editorial  Expanders - Apply equivalent subjects  Search modes - Boolean/Phrase | 28 |

^1^ These databases were simultaneously accessed through EBSCOhost.

eMethods. The effects of megadrivers on health.

Globalisation

In his new book, The Ages of Globalisation: Geography, Technology, and Institutions, renowned economist Jeffrey Sachs outlines a series of seven distinct waves of technological and institutional change (starting with the original settling of the planet by early modern humans through long-distance migration and ending with reflections on today’s globalisation) [1]. He argues that the historical interplay of geography, technology, and institutions has impacted the ongoing processes taking place today: globalisation based on digital technologies. He emphasises the need for new methods of international governance and cooperation to prevent conflicts and to achieve economic, social, and environmental objectives aligned with sustainable development.

Globalisation has had various positive and negative implications on health [2]. While the interconnectedness of globalisation has afforded the global sharing of ideas, enjoyment of diverse cultures, and trade of goods, recent pandemics have proven that diseases know no borders and with increasing globalisation, diseases can spread and are spreading more rapidly than ever before. This interconnectedness can, however, also be a part of the solution. The current COVID-19 pandemic has shown how a coordinated global effort can lead to swift responses to vaccine development.

Digitalisation

Digitalisation of health (with digital health as its outcome) has had an escalating impact on health, healthcare, living, and society, presenting commensurate challenges and opportunities for human progress. In The Fourth Wave: Digital Health a New Era of Human Progress, Paul Sonnier explains how the fusion of the digital and genomic revolutions is rapidly creating a new era of human progress, spurred by specific key drivers and facilitated by megatrends [3]. He highlights various opportunities of digital health, such as combating chronic diseases (e.g. stimulating activity to address a common risk factor of cardiovascular disease), improved diagnostic processes through genetic testing or improved imaging, and novel healthcare options through gene therapy (e.g in certain forms of leukemia or certain forms of blindness) or human augmentation (e.g. exoskeletons for people that suffered a stroke).

Increasing digitalisation and digital transformations have not only revolutionised how we interact with our health and the environment, but also have highlighted their potential to contribute to the development of potential solutions for the wicked problems mapped in this study. In certain contexts, they have even exacerbated underlying determinants of health and contributed to the widening of the existing digital divide. Of those left disconnected as a result of increasing digitalisation and digital transformations, many are disproportionately affected (specific population subgroups include: youth (aged 15-24; [4]), women and girls, gender and sexual minorities, marginalised races, religious and ethnic groups, the homeless, refugees, and other underrepresented communities) [5]. On the other hand, those who remain connected may face more online risks as a result of increased digital (media) exposure [6].

The recently established The Lancet and Financial Times “Governing Health Futures 2030: Growing up in a digital world” Commission has recently been established to explore the convergence of digital health, artificial intelligence (AI) and other frontier technologies with UHC to support attainment of the third Sustainable Development Goal (SDG), and provide recommendations for policymakers on the governance of (digital) health futures [7].

As children and young people are growing up in an increasingly digital world, digital transformations present opportunities and threats for adolescent health and well-being (Holly, Wong, et al., 2021 [forthcoming])—in particular, across five domains of adolescent well-being: connectedness, positive values, and contribution to society; good health and optimum nutrition; safety and a supportive environment; learning, competence, education, skills, and employability; and agency and resilience [8].

Demographic Change

Demographic change is an umbrella term and includes a number of factors, such as ageing, declining fertility rates, and diverse migration flows [9,10]. While considerable effort has been put in understanding the effects of ageing on the functioning and financing of the healthcare system in particular, the effects of demographic change on health are far wider [9]. In terms of an ageing population, sexual and mental health are becoming increasingly important as the concept of healthy ageing gains traction in public health strategies [9,11]. Additionally, ageing is found to have explicit, yet divergent effects on the burden of disease of some communicable diseases [12]. In terms of migration flows, the influx of migrants from Africa and the Middle-East combined with the continuing exodus of the health workforce towards Western Europe remains an exigent public health priority [9].

The United Nations has proclaimed 2021–2030 the “Decade of Healthy Ageing”, with the WHO leading international action to improve the lives of older people, their families, and communities. In doing so, the Decade convenes a variety of stakeholders with concerted action to: (1) change how we think, feel, and act towards age and ageing; (2) develop communities in ways that foster the abilities of older people; (3) deliver person-centred, integrated care, and primary health services that are responsive to older people; and (4) provide older people access to long-term care when they need it [11]. While an ageing demographic worldwide has implications on health systems and policies, these extend to youth who will be ageing within these systems/structures (Wong, Rangan, et al., 2021; forthcoming) [ref].
